# Data-driven characterization of individuals with delayed autism diagnosis

**DOI:** 10.1101/2024.07.26.24311003

**Authors:** Dan Aizenberg, Ido Shalev, Florina Uzefovsky, Alal Eran

## Abstract

**Importance:** Despite tremendous improvement in early identification of autism, ∼25% of children receive their diagnosis after the age of six. Since evidence-based practices are more effective when started early, delayed diagnosis prevents many children from receiving optimal support.

**Objective:** To identify and comparatively characterize groups of individuals diagnosed with Autism Spectrum Disorder (ASD) after the age of six.

**Design:** This cross-sectional study used various machine learning approaches to classify, characterize, and compare individuals from the Simons Foundation Powering Autism Research for Knowledge (SPARK) cohort, recruited between 2015-2020.

**Setting:** Analyses of medical histories and behavioral instruments.

**Participants:** 23,632 SPARK participants.

**Exposure:** ASD diagnosis upon registration to SPARK.

**Main Outcomes and Measures:** Clusters of individuals diagnosed after the age of six (*delayed ASD diagnosis*) and their defining characteristics, as compared to individuals diagnosed before the age of six (*timely ASD diagnosis*). Odds and mean ratios were used for feature comparisons. Shapley values were used to assess the predictive value of these features, and correlation-based cliques were used to understand their interconnectedness.

**Results:** Two robust subgroups of individuals with delayed ASD diagnosis were detected. The first, *D1*, included 3,612 individuals with lower support needs as compared to 17,992 individuals with a timely diagnosis. The second subgroup, *D2*, included 2,028 individuals with higher support needs, as consistently reflected by all commonly-used behavioral instruments, the greatest being repetitive and restrictive behaviors measured by the Repetitive Behavior Scale – Revised (RBS-R; D1: MR = 0.6854, 95% CI = 0.6848 – 0.686; D2: MR = 1.4223, 95% CI = 1.4210-1.4238, P = 3.54 × 10^−134^). Moreover, individuals belonging to D1 had fewer comorbidities as compared to individuals with a timely ASD diagnosis, while D2 individuals had more (D1: mean = 3.47, t = 15.21; D2: mean = 8.12, t = 48.26, p< 2.23 × 10^−308^). A Random Forest classifier trained on the groups’ characteristics achieved an AUC of 0.94. Further connectivity analysis of the groups’ most informative characteristics demonstrated their distinct topological differences.

**Conclusions and Relevance:** This analysis identified two opposite groups of individuals with delayed ASD diagnosis, thereby providing valuable insights for the development of targeted diagnostic strategies.

**Key Points:** *Question:* Are there specific subgroups of individuals diagnosed with autism after school age?

*Findings:* In this data-driven analysis of a large cohort of autistic individuals, two distinct subgroups of individuals diagnosed with autism after the age of six were identified. The first included individuals requiring low levels of support, with modest comorbidity burdens; The second included individuals requiring high levels of support, with extremely high comorbidity burdens.

*Meaning:* The identification of opposite subgroups of individuals with delayed autism diagnosis improves our understanding of autism heterogeneity and moves us closer towards precision diagnosis of autism.

## Introduction

One in 36 children in the United States is diagnosed with Autism Spectrum Disorder (ASD), a heterogeneous spectrum of neurodevelopmental conditions characterized by difficulties in social cognition and communication, and restrictive and repetitive behavior (RRBs)^1^. Early intervention, starting as early as 12 months of age, can substantially affect the development and long-term outcomes of autistic children^2–6^. However, despite dramatic improvements in the proportion of children who receive a developmental screening by age 36 months, the median age of ASD diagnosis in the US is currently 49 months. Thus, many autistic individuals remain undiagnosed until and throughout school-age^1,7,8^, preventing them from receiving the support they need.

Delayed diagnosis can be partially explained by demographic factors, including sex, race^9–11^, socio-economic status^12,13^, and autism awareness at the time of diagnosis^13^. Delayed diagnosis may also be attributed to the severity of autistic traits and one’s level of required support^9–11^. However, these factors explain delayed diagnosis only partially, and most of the variance in the age of ASD diagnosis remains unexplained^12–14^. Previous studies investigating delayed ASD diagnosis have typically focused on a limited set of characteristics^12,13,15,16^. Moreover, these studies have treated individuals with delayed diagnosis as a single group, while autism is highly heterogeneous^17,18^.

Autistic children often suffer from co-occurring conditions, including seizures, gastrointestinal dysfunction, and growth problems^19–30^. Some of these conditions have been shown to share genetic components with autism^31–33^, correlate with other common autism comorbidities^34^, or be associated with autistic traits^35–37^. An association between some comorbidities and delayed diagnosis might arise from several reasons. The first is overshadowing of autism by a co-occurring disorder, or in other words, attributing autistic traits to another disorder that may or may not co-occur with ASD^22,38,39^. For example, ASD may be overshadowed by attention deficit hyperactivity disorder (ADHD), and it has been shown that a presentation of hyperactivity or inattention often leads to a primary diagnosis of ADHD, consequently delaying one’s autism diagnosis^33,40^. Moreover, a set of comorbidities may present a unique profile that deflects the diagnostic course from ASD.

Another potential reason for certain comorbidities to be associated with delayed ASD diagnosis is that such comorbidities might share an underlying genetic basis with autistic traits appearing later in life^34^. For example, that might be the case of phenylketonuria^41^ or Smith-Lemli-Opitz syndrome (SLOS)^42^. Therefore, a comprehensive characterization or profiling of individuals with delayed ASD diagnosis using their phenotypic and comorbidity profiles may shed light on the underlying processes associated with delayed diagnosis, ultimately enabling targeted screening options and improved outcomes.

Recently, attempts have been made to exploit machine learning (ML) and big data to dissect the heterogeneous autism spectrum, based, for example, on behavioral phenotypes^43^, neuroanatomy and neuro functioning^44,45^, information processing ability^45^, and comorbidity profiles^35–37,46^. However, to date, no data-driven approach has been used to dissect delayed diagnosis. Such a large-scale systematic approach could be used to explain diagnostic delays and gain a better understanding of the heterogeneity of autism. This, in turn, could enable targeted research and diagnosis options.

Here we analyzed a large cohort of autistic individuals, focusing on those diagnosed after the age of six, considered as having a delayed diagnosis. Using various supervised and unsupervised ML algorithms, we identified and characterized two distinct groups of individuals with delayed diagnosis (denoted D1 and D2). We also leveraged this big data to quantify the relationships between the characteristics of each group.

## Methods

The Boston Children’s Hospital Institutional Review Board has determined that this research qualifies as exempt from the requirements of human subjects protection regulations.

### Data preprocessing

We mined the SPARK phenotype dataset (version 5)^45^, containing rich phenotypic data from 99,447 autistic individuals, aged 0-92 years. Tables populated with 25% or more of the data were used for our analysis. These include basic medical screening, Social Communication Questionnaire-Lifetime (SCQ)^47^, Background history questionnaire, Repetitive Behavior Scale-Revised (RBS-R)^48^, and Developmental Coordination Disorder Questionnaire (DCDQ)^49^. Of these tables, only relevant features were considered (**eTable 1**). Of note, IQ and familial medical and mental health history were not collected on a sufficiently large population and were therefore excluded from the analysis. Continuous variables were Z-standardized, and categorical variables were represented as one-hot vectors. All data was processed using Python’s pandas v1.3.4^50^ and NumPy v1.20.3^51^ packages.

A total of 23,632 individuals who had complete information in all selected tables and whose autism diagnosis was never refuted or remitted (N = 1727) were included in the analysis. Their median age of diagnosis was 3.67 years (IQR = 2.58 – 5.75 years).

### Definition of delayed diagnosis

The age cutoff for delayed diagnosis was six years, chosen for being higher than the 75^th^ percentile of diagnosis ages in the studied cohort (5.75 years). Moreover, six years of age was previously reported to be a key turning point in autism, in which the course of autistic traits and behavior sometimes dramatically changes^52^. Autistic traits and behavior appear much earlier than this age, and by then, the relevant time window for early intervention has long since passed^5,53^. Accordingly, participants were first divided into one of two groups: those diagnosed before the age of six (median = 3.08 years, IQR = 2.42 – 4.08 years), and those diagnosed at or after the age of six (median = 8.00 years, IQR = 6.83 – 9.67 years). **eFigure 1** shows the number of individuals in each group and the distribution of their ages at diagnosis, birth year, and year of diagnosis.

### Identification of groups of individuals with delayed ASD diagnosis

K-means clustering of individuals diagnosed at or after the age of six was performed on all standardized variables listed in **eTable 1**, using Python’s scikit-learn package v0.23.2^54^. The number of clusters was identified using the elbow method, ensuring that every cluster contained at least 10% of individuals.

### Group characterization

To characterize the delayed diagnosis groups, each variable listed in **eTable 1** was tested for differences between its distribution in each group and 1,000 bootstrapped samples from the timely diagnosis group, with the bootstrap sample size equal to the group size. For binary variables, Fisher’s exact tests were performed between these samples. For continuous variables, Mann–Whitney U tests were performed and the ratio of the groups’ means was saved as an interpretable statistic comparable to the odds-ratio statistic from the Fisher’s exact test. Benjamini– Hochberg correction was then used to account for multiple testing, ensuring a false discovery rate of 0.05. Python’s pandas v1.3.4^50^ was used for bootstrapping, SciPy stats^55^ v1.7.1 was used for Fisher’s and Mann-Whitney tests, and Statmodels^56^ v0.12.1 was used for their corrections.

### Analyzing relations between co-occurring psychiatric disorders and age of ASD diagnosis

Pearson correlations were calculated between the age of ASD diagnosis and the prevalence of reported psychiatric disorders, using SciPy^55^ stats v1.7.1. Benjamini–Hochberg corrections were used to account for multiple testing (α = 0.05, Statmodels^56^ v0.12.1).

### Comorbidity burden comparison

For each individual, we summed the number of reported comorbid conditions. Pairwise two-sample t-tests were used to compare these between identified using SciPy^55^ v1.7.1.

### Group classification and model inference

A random forest classifier was trained to classify individuals into one of identified groups using Python’s scikit-learn^54^ package v0.23.2, on a training set of 75% of the data. Underrepresented clusters were synthetically over-sampled in the training set using the SMOTE-NC algorithm^57^, implemented in Python’s imbalanced-learn^58^ package v0.7.0. The classifier’s hyper-parameters were tuned using a grid-search with 10-fold cross-validation and with micro-averaged F1-score as the scoring method. To understand the contribution of features to the classification, Shapley values were calculated using Python’s SHAP package, v0.37.0^59^.

### Feature interconnectivity and network analysis

To understand the relations between features of each group, we calculated Pearson’s correlations between each pair of features in each group. We represented these relations in a graph whose feature vertices were joined by an edge if their correlation coefficient was > 0.1 and their Benjamini-Hochberg adjusted correlation P value was < 1×10^-4^. We then identified correlation-based cliques using Python’s NetworkX v2.6.3^60^. We specifically focused on 4-cliques, defined as groups of four or more vertices that are all connected to each other. Normalized clique membership was determined by dividing the number of 4-cliques in which each (vertex) variable appeared within a group by the total number of cliques in that group.

## Results

### Two distinct groups of individuals with delayed ASD diagnosis differ by their co-occurring conditions and levels of autistic traits

In all, 5,640 of 23,632 (23.87%) SPARK participants received their ASD diagnosis after the age of six, consistent with recent US-wide Autism and Developmental Disabilities Monitoring (ADDM) network reports^61^. K-means clustering revealed two distinct groups of autistic individuals diagnosed after the age of six (**eFigure 2**). The first, D1, included 3,612 individuals (64%), and the second, D2, included 2,028 individuals (36%). D1 individuals were characterized by fewer autistic traits than timely diagnosed individuals, whereas D2 individuals had more autistic traits (**Figure 1**). This finding is consistent across all autism domains, as measured by multiple behavioral instruments, and spans difficulties in social communication and RRBs (**Figure 1A**), motor development (**Figure 1B**), and feeding and sleeping problems (**Figure 1C**).

**Figure 1.**
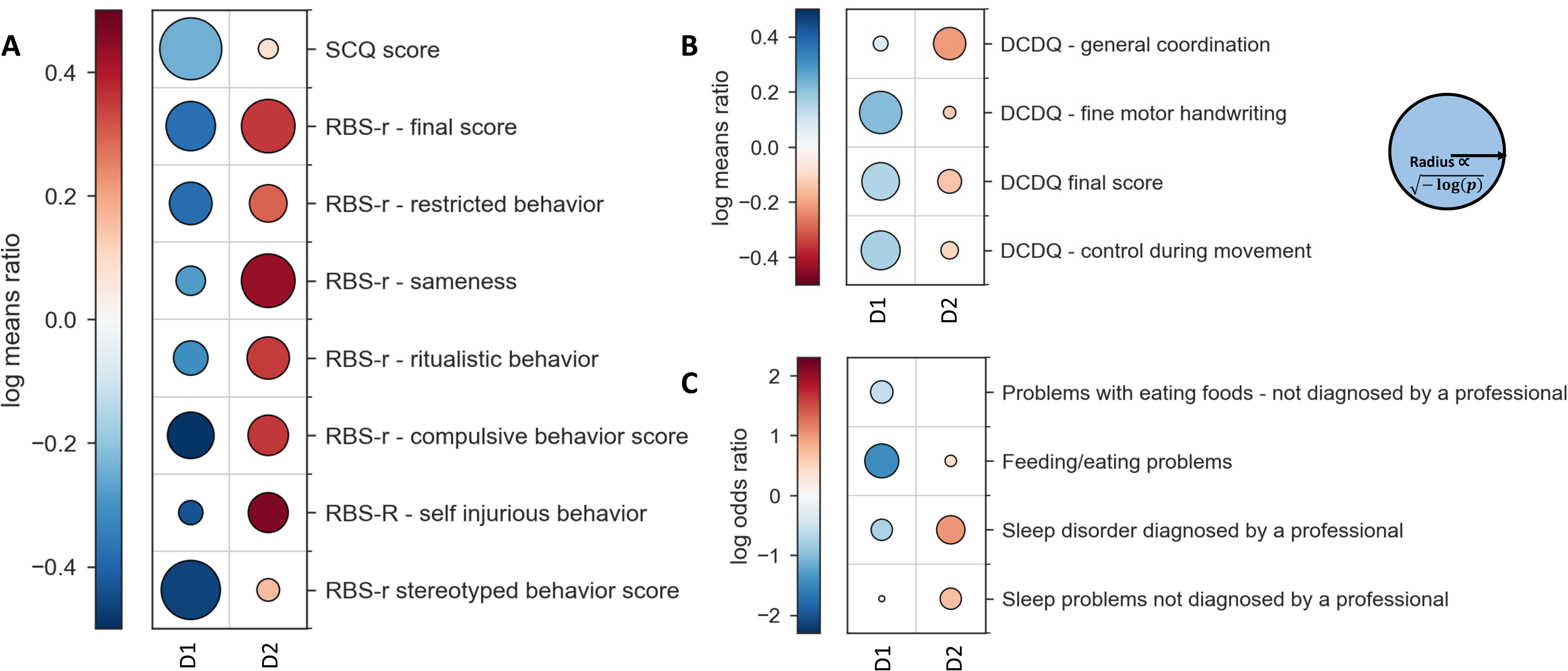
Behavioral differences between groups of individuals with delayed ASD diagnosis. Differences between groups of individuals with delayed ASD diagnosis (D1, D2) and those with a timely one, in (**A**) core autism domains, (**B**) motor development, and (**C**) common symptoms. The color of each circle represents the effect size measured by the log ratio of a feature’s mean in each group as compared to the feature’s mean among timely diagnosed individuals. The size of the circles is proportional to 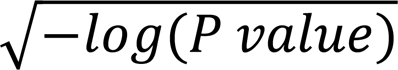 . Thus, the larger the circle, the smaller the P value. The lowest possible P value in this analysis is 6. 87 × 10^−278^. Circles are shown only for significant results.

Moreover, D1 individuals had lower odds of co-occurring neurodevelopmental conditions than timely diagnosed individuals, whereas D2 individuals had higher odds for such conditions (**Figure 2A-C**). These include cognitive and motor delays (**Figure 2A**), growth abnormalities (**Figure 2B**), and neural problems (**Figure 2C**). While both delayed diagnosis groups had higher rates of ADHD, anxiety, and depression or dysthymia as compared to timely diagnosed individuals, those in D2 had higher rates of neuropsychiatric disorders, including obsessive-compulsive disorder (OCD), oppositional defiant disorder (ODD), and schizophrenia (**Figure 2D**). Moreover, a positive linear trend was found between the age of diagnosis and the prevalence of neuropsychiatric disorders (**eFigure 3**).

**Figure 2.**
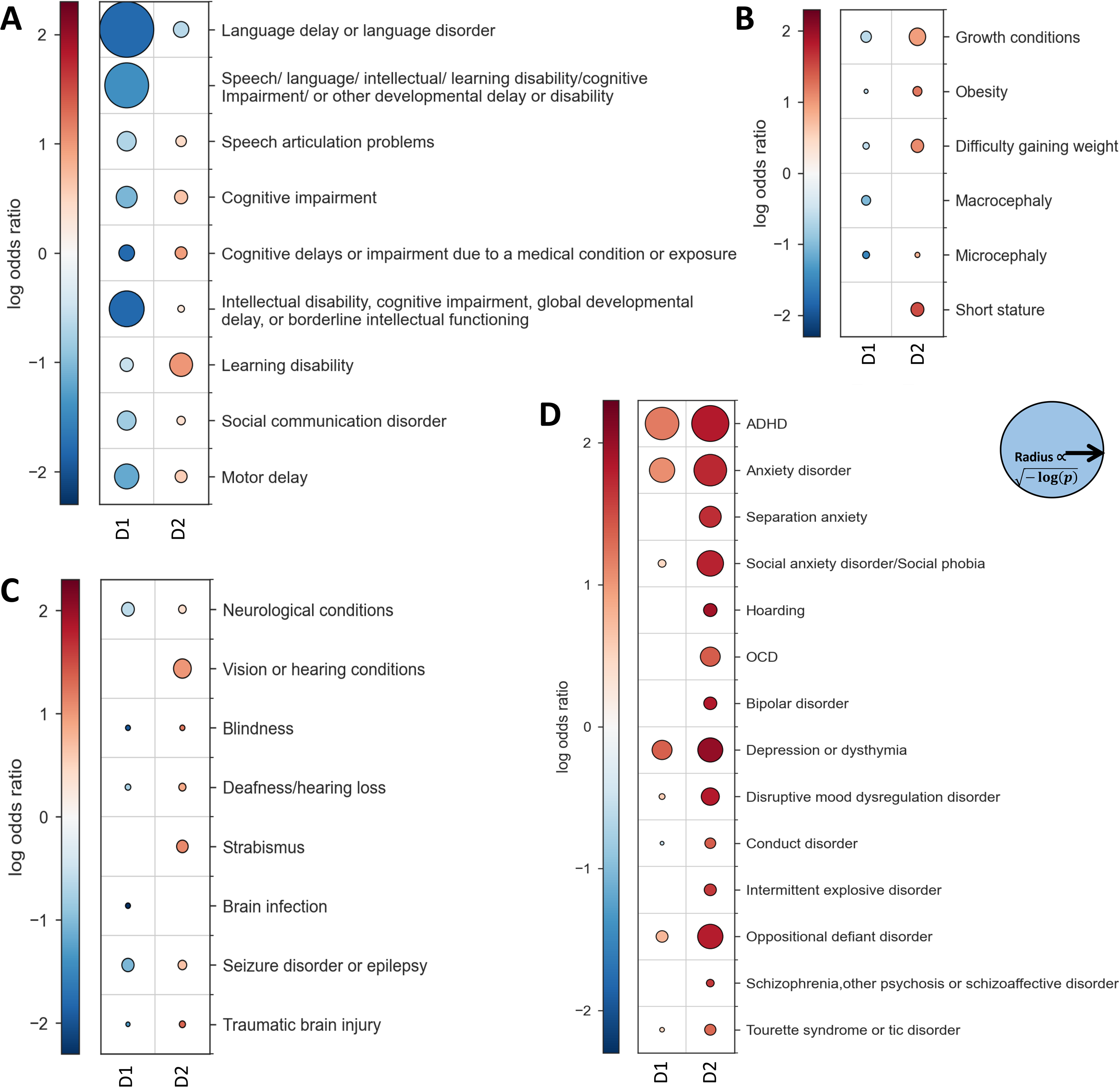
Differences in co-occurring neurodevelopmental conditions between groups of individuals with delayed ASD diagnosis. (**A**) Differences in cognitive and motor delays; (**B**) differences in growth alterations; (**C**) Differences in sensory and neural problems; and **(D)** Differences in neuropsychiatric conditions. The color of each circle represents the effect size measured by the log ratio of a feature’s mean in each group as compared to the feature’s mean among timely diagnosed individuals. The size of the circles is proportional to 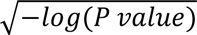. Thus, the larger the circle, the smaller the P value. The lowest possible P value in this analysis is 6. 87 × 10^−278^. Circles are shown only for significant results.

Additionally, D1 individuals had lower odds of congenital disabilities than timely diagnosed individuals, whereas D2 individuals had higher odds for such conditions (**eFigure 4**). Similarly, prenatal and perinatal complications were less common in individuals in the D1 group as compared to timely diagnosed individuals, whereas individuals in the D2 group had higher odds for such conditions, including fetal alcohol syndrome, premature birth, and insufficient oxygen at birth (**eFigure 5**). Finally, parents of individuals in D1 were more educated than parents of timely diagnosed individuals, while parents of individuals in D2 were less educated (**eFigure 6**).

### Groups of individuals with delayed diagnosis are characterized by extreme rates of co-occurring conditions

Next, we compared the total number of co-occurring conditions between groups. While individuals with a timely ASD diagnosis had, on average, 4.34 co-occurring conditions, D1 individuals had significantly less (mean = 3.47, t = 15.21, p = 3.01 × 10^−52^) and D2 individuals had significantly more comorbidities (mean = 8.12, t = 48.26, p < 2.23 × 10^−308^; **Figure 3**). Thus, individuals with delayed ASD diagnosis are characterized by extreme rates of co-occurring conditions.

**Figure 3.**
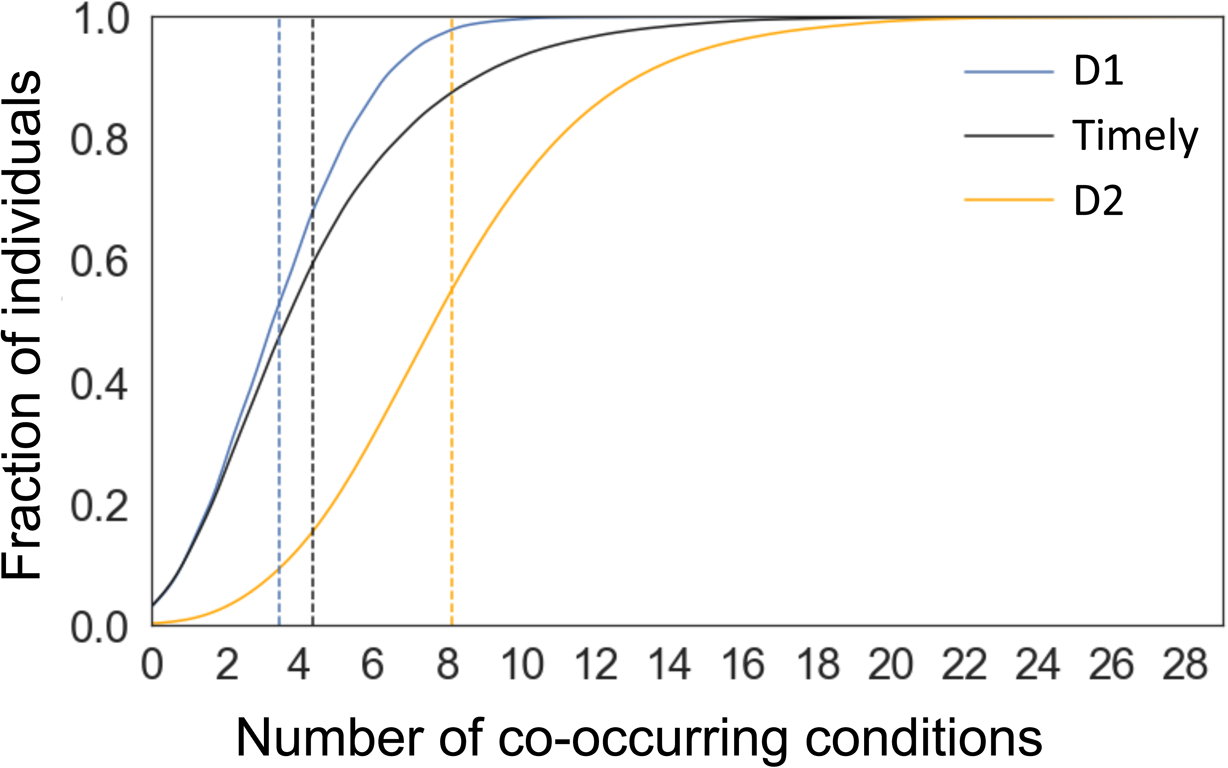
Cumulative distributions of co-occurring conditions. Cumulative distributions of the number of co-occurring conditions in individuals of each group. Horizontal lines depict the mean number of co-occurring conditions in each group.

### Levels of autistic traits and prevalence of psychiatric disorders are the primary distinguishing features between delayed diagnosis groups

We next determined how distinguishable are the two groups of individuals with delayed diagnosis and what features contribute most to their classification. We trained a one vs. rest random forest classifier to assign an autistic individual to one of the identified groups. The model achieved a micro-averaged AUC of 0.94 and a micro-averaged average-precision of 0.89 on a test set of 25% of the data (**eFigure 7**). Thus, the identified groups could be distinguished using a relatively simple model. The most important features in this model were identified by their Shapley values, i.e., their relative contribution to the model, considering all possible combinations of the remaining features.

Of the 20 most important features distinguishing D1 from D2 and the timely diagnosis group, 14 overlap and predict in opposite directions, reflecting the two extremes of the delayed diagnosis groups (**Figure 4**). These features include language disorders, RBS-r subscales, SCQ scores, learning disorders, sleeping problems, DCDQ scores, and DCDQ’s fine motor control subscale score. Moreover, mood disorders, anxiety disorders, depression, or OCD were found to be informative characteristics of D2. Contrarily, lack of intellectual disability, speech articulation problems, or problems with eating foods contributed to the assignment of an individual to D1.

**Figure 4.**
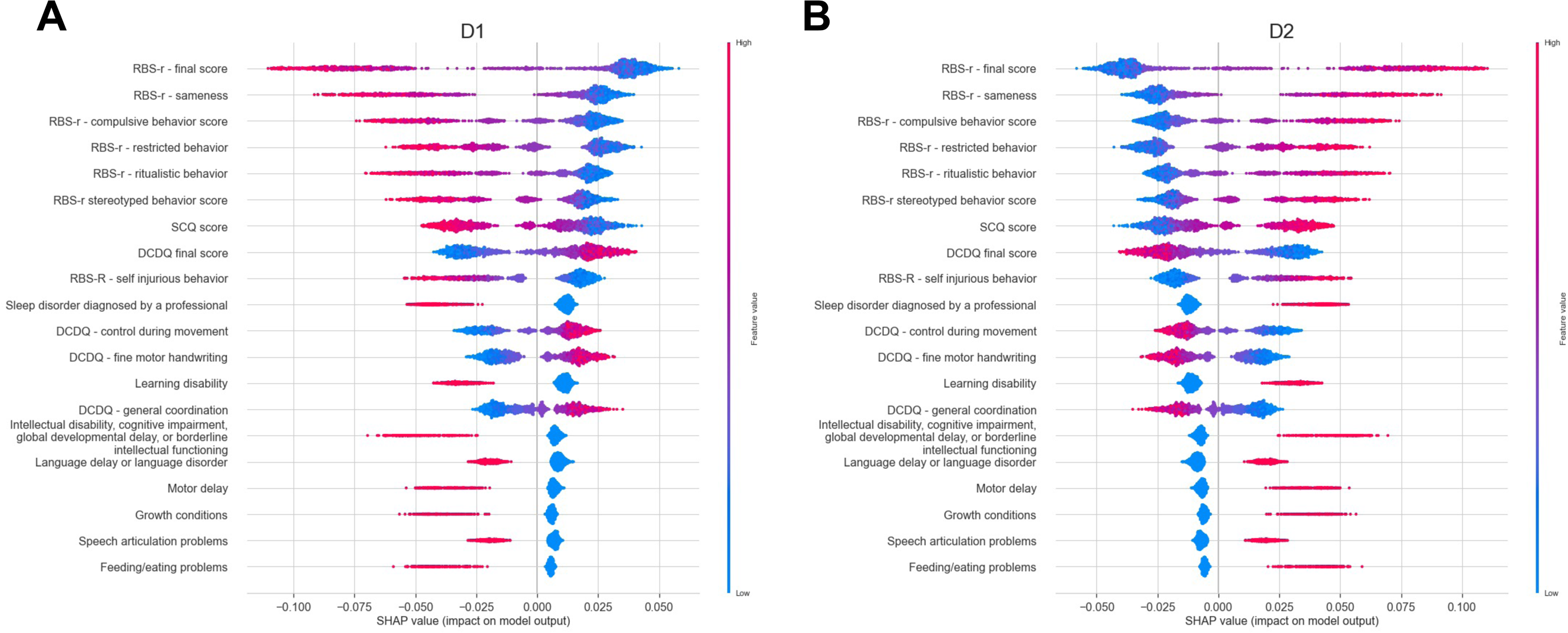
Feature importance using Shapley values. Most distinguishing features of (**A**) D1, and (**B**) D2, based on Shapley values. Each dot in the plot corresponds to a sample. The features are ordered on the y-axis by their total impact on the model output. The x-axis shows the relative impact on the model output for a given sample. The color shows the relative sample value for every feature.

### Relationships between group characteristics demonstrate unique relations between autistic traits and co-occurring conditions

Finally, we examined inter-relations between the most informative characteristics of each group. Toward that end, we calculated pairwise correlations among D1 features (**eTable 2**), D2 features (**eTable 3**), and those of the timely diagnosis group (**eTable 4**). We then assessed the strength of information flow inherent to each feature by modeling these correlations as a graph and calculating each feature’s normalized clique membership, which indicates the percent of 4-cliques that contain that feature (**eTable 5**). The larger a feature’s normalized clique membership, the more information flows through it. **Figure 5A-C** depicts and compares the five most inter-connected features in each group, demonstrating the difference between the role these features play in each group. For example, in D1, the two most inter-connected variables are related to RRBs, and in D2 it is motor delay and birth disabilities. To gain a more comprehensive understanding of inter-relations between the most connected features within each group, correlation heatmaps were used to depict pairwise correlations between the most connected features and their clique members in D1 (**Figure 5D**), D2 (**Figure 5E**), and the timely diagnosed group T (**eFigure 8)**. These heatmaps demonstrate group-specific topology of feature relatedness.

**Figure 5.**
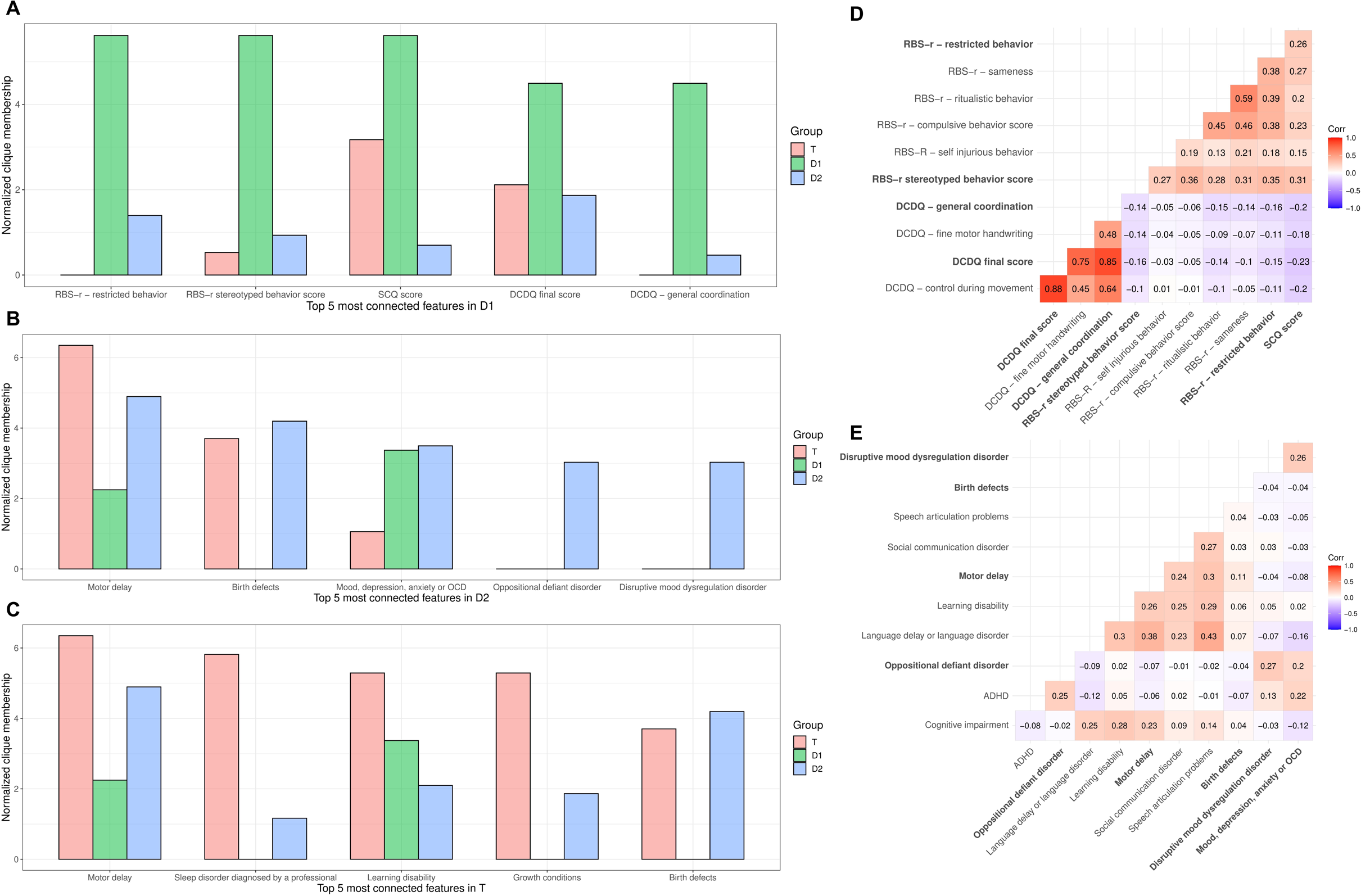
Feature interconnectivity and network structure of timely and delayed diagnosis groups. Normalized clique membership is depicted for the top five most connected features in (**A**) D1, (**B**) D2, and (**C**) the timely diagnosis group, T. Correlation heatmaps of the variables most strongly correlated with the top connected features are presented for (**D**) D1, and (**E**) D2. The top connected features are denoted in bold.

## Discussion

Tremendous efforts are being made worldwide to lower the age of ASD diagnosis, since early detection can greatly improve outcomes and intervention opportunities^2–6^. The present study leveraged novel machine learning approaches and big data from SPARK to identify factors contributing to delayed diagnosis. We identified two distinct groups of autistic individuals with delayed diagnosis, differing in the level of autistic traits and the degree of co-occurring conditions, and displaying distinct network structures.

Autism is highly heterogenous, and as such, a single reason for late diagnosis is unlikely. Yet, previous studies examining the differences between late and early diagnosed individuals lacked the design or analytical power to untangle such heterogeneity, resulting in conflicting evidence. While some studies suggested higher rates of co-occurring conditions^62^ and no differences in autistic traits in the late-diagnosed group^15^, others reported more pronounced autistic traits and higher rates of neuropsychiatric conditions, together with communication difficulties^63^, language delay^13^, and intellectual disability^9,13^ as factors enabling earlier diagnosis. Our findings suggest the existence of two mirroring groups of individuals with delayed ASD diagnosis, which may explain the conflicting results observed in previous studies.

The first identified group, D1, was characterized by lower levels of autistic behaviors and lower odds of co-occurring neurodevelopmental and neuropsychiatric conditions, as compared to the timely diagnosed group. Individuals in this group also displayed fewer learning and intellectual disabilities and better verbal skills. These characteristics align with previous findings showing that these characteristics contribute to delayed autism diagnosis^9,12,15,64,65^. It is possible that higher verbal and intellectual skills may delay parents in seeking a diagnosis or being referred to dedicated clinics, thus delaying the time of diagnosis.

Contrarily, the second group, D2, showed higher levels of autistic traits and high rates of neurodevelopmental disorders or delays. In this group, autism is only one of many co-occurring conditions. Autism evaluations are typically focused on the specific clinical features of autism, ignoring co-occurring conditions, and could be overshadowed by them^66^. Consequently, the high rates of co-occurring conditions could make the recognition of autism more complex, delaying the age of diagnosis.

The two groups were further distinguished based on their underlying network structure and feature interconnectivity. RRBs were identified as the core characteristic of D1, showing tight intercorrelations within this group. Recent findings involving younger children with a strong indication for ASD (suggesting timely diagnosis), showed strong connectivity among RRBs, but it was considered a peripheral dimension of the network in that sample, with social communication difficulties identified as the core aspect of the network^67^. In contrast, our analysis suggests that for some individuals with delayed diagnosis, RRBs and their connectivity represent the core feature of autism.

Network analysis identified different core features in D2, such as motor delay and congenital disabilities. These conditions correlated with difficulties in social communication, reinforcing the idea that co-occurring conditions might overshadow ASD diagnosis^66^. Nevertheless, further research is needed to explore the interplay between characteristics within these groups.

Besides offering targeted approaches for future screening and diagnosis, this study can help make sense of the heterogeneity characterizing autism. Such heterogeneity is reflected by complex genetic etiology with over one thousand known genetic variants associated with autism, most with small effect sizes and additive effects^18,68^. The two groups identified in our study may reflect differences in physiological and phenotypic bases that could be used for future targeted genetic research and better characterization of autism.

This study holds several limitations. Key among them is the absence of temporal data, such as the onset of co-occurring mental and developmental conditions, limiting our ability to clarify if and how these factors precede ASD diagnosis. Previous research suggests that preceding neurodevelopmental and neuropsychiatric conditions contribute to delayed ASD diagnosis^40,62^. For example, children with a prior ADHD diagnosis, tend to receive their ASD diagnosis an average 1.8 years later than children without ADHD^40^. Future research should employ longitudinal designs to investigate the temporal directionality of D1- and D2- informative characteristics identified in our study.

Other limitations include the exclusion of IQ measures from our data due to missing data, which could have provided valuable insights. Furthermore, SPARK participants are mostly White, limiting the generalizability of our study, and future studies should include a more diverse cohort. Finally, throughout the paper, we consistently used the term ‘ASD’ (or individuals with an ASD diagnosis), to refer to the official diagnosis given to participants according to the Diagnostic and Statistical Manual of Mental Disorder (DSM). However, we acknowledge the controversy surrounding the use of this term, and when referring to individuals, we used identity-first language to describe autistic people, as it is the preferred choice of most autistic individuals^69^.

## Conclusions

In this study, we identified and characterized two distinct groups of individuals diagnosed with ASD after the age of six. By dissecting the heterogeneous landscape of individuals with delayed ASD diagnosis, this work opens new avenues for better-powered (less heterogenous) genetic and behavioral research. Ultimately, this work enables the development of targeted diagnostic strategies and thereby improved outcomes for autistic children.

## Supporting information

Supplementary online content

## Data Availability

All data produced in the present work are contained in the manuscript.

## Author contributions

The original draft of the paper was written by DA, who also conducted all the main statistical analyses. DA, IS, FU, and AE were responsible for both the design of the study and the interpretation of its results. The original draft was reviewed and edited by AE, FU, and IS, who made critical contributions to the paper’s content. IS performed some of the secondary analyses. The project was supervised by AE and FU, who provided funding for this study. All authors approved the submitted version of this paper. AE had full access to all of the data in the study and takes responsibility for the integrity of the data and the accuracy of the data analysis.

## Additional Contributions

We are grateful to all of the families in SPARK, the SPARK clinical sites and SPARK staff. We appreciate obtaining access to SPARK phenotypic collection (version 5) data on SFARI Base. Approved researchers can obtain the SPARK population dataset described in this study ([SPARK Phenotype Dataset(https://www.sfari.org/resource/spark/) by applying at https://base.sfari.org. Additionally, we would like to thank the Uzefovsky and Eran lab members for engaging in valuable and fruitful discussions.

