## Supplementary online content for "Data-driven characterization of individuals with delayed autism diagnosis"

**eTable 1.** SPARK variables included in the analysis.

**eTable 2.** Pairwise correlations between informative characteristics of the D1 group.

**eTable 3.** Pairwise correlations between informative characteristics of the D2 group.

**eTable 4.** Pairwise correlations between informative characteristics of the timely diagnosis group.

**eTable 5.** Feature participation in pairwise correlation 4-cliques.

**eFigure 1.** Sex and age of autism diagnosis of SPARK individuals included in this study.

**eFigure 2.** Two groups of individuals with delayed autism diagnosis.

**eFigure 3.** Prevalence of psychiatric disorders according to the age of autism diagnosis.

**eFigure 4**. Differences in congenital disorders between individuals in each delayed diagnosis group and those that received a timely diagnosis.

**eFigure 5**. Differences in prenatal and perinatal complications between individuals in each delayed diagnosis group and those with a timely diagnosis.

**eFigure 6**. Differences in family and demographics between individuals in each delayed diagnosis group and those with a timely autism diagnosis.

**eFigure 7.** Receiver operating characteristics (ROC) curve for distinguishing the two delayed diagnosis groups using a random forest classifier.

**eFigure 8.** Heatmap of variables most strongly correlated with the top connected features of the timely diagnosis group.

**eTable 1. SPARK variables included in the analysis.**

| Included variables |
|---|
| ADHD (Attention Deficit-Hyperactivity Disorder) or ADD |
| Age of diagnosis in months: how old was child/dependent when he/she was first diagnosed |
| Alcohol or Substance Use |
| Annual household income |
| Annual household income (if parents are divorced, report income at the child's primary residence): |
| Anxiety disorder, such as panic, phobia, agoraphobia, or generalized anxiety disorder (GAD) except for social anxiety |
| Attention or behavior disorders |
| Bipolar (Manic-Depressive) Disorder |
| Birth defects |
| Birth defects of bones, hands or feet |
| Birth or pregnancy complications |
| Bleed into the brain |
| Blindness |
| Brain and spinal cord birth defects |
| Brain infection such as bacterial meningitis, encephalitis |
| Brain malformation/abnormality (shown on MRI) |
| Brother with ASD |
| Cataract |
| Child with ASD |
| Cleft lip |
| Cleft palate |
| Clubbed foot |
| Cognitive delays or impairment due to another medical condition or exposure (For example, brain injury, stroke, lead poisoning, FAS, HIV, radiation, hydrocephalus, brain tumor, drug effects, etc.) |
| Conduct Disorder |
| Congenital diaphragmatic hernia |
| Congenital heart disease/defect |
| Control during movement subscale score (DCDQ) |
| DCDQ total score |
| Deafness/hearing loss |
| Depression or dysthymia |
| Difficulty gaining weight |
| Disruptive Mood Dysregulation Disorder |
| Eating Disorder |
| Esophageal atresia (no connection between the esophagus and stomach) |
| Extra fingers and/or extra toes |
| Facial birth defect |
| Father with ASD |
| Father's biological brother or sister with ASD |
| Feeding/eating problems |

| |
|---|
| Fetal Alcohol Syndrome, alcohol or drug exposure in mother's pregnancy |
| Fine Motor/Handwriting subscale score (DCDQ) |
| Gastrointestinal (GI) birth defects |
| General Coordination subscale score (DCDQ) |
| Growth conditions (including height, weight and head size) |
| Heart or lung birth defect |
| Highest level of education of father/guardian: |
| Highest level of education of mother /guardian: |
| Highest level of education of mother/guardian: |
| Hirschsprung disease |
| Hoarding |
| How many weeks (gestational age) was child/dependent when he/she was born? |
| Hypospadias (in boys, urinary opening is in the wrong place) |
| I. Stereotyped Behavior- Total Subscale Score (RBS-r) |
| II. Self-Injurious Behavior- Total Subscale Score (RBS-r) |
| III. Compulsive Behavior- Total Subscale Score (RBS-r) |
| Insufficient oxygen at birth with NICU stay |
| Intellectual disability or cognitive impairment |
| Intellectual disability, cognitive impairment, global developmental delay, or borderline intellectual functioning |
| Intermittent Explosive Disorder |
| Intestinal malrotation |
| IV. Ritualistic Behavior- Total Subscale Score (RBS-r) |
| Kidney malformation (for example horseshoe kidney) |
| Language delay or language disorder |
| Large head size (macrocephaly) |
| Lead poisoning |
| Learning disability (LD, learning disorder, including reading, written expression, math, or NVLD (Nonverbal learning disability)) |
| Lung malformation |
| Missing kidney |
| Missing or malformed bones |
| Missing uterus |
| Monozygotic twin |
| Mood, Depression, Anxiety or OCD |
| Mother with ASD |
| Mother's biological brother or sister with ASD |
| Motor delay (e.g., delay in walking) or developmental coordination disorder |
| Multiple birth |
| Mutism |
| Neurological conditions |
| No family member with ASD |
| Obesity |
| Obsessive-Compulsive Disorder |
| One of cousins on father's side with ASD |
| One of cousins on mother's side with ASD |

| |
|---|
| One of grandparents on father's side with ASD |
| One of grandparents on mother's side with ASD |
| Oppositional Defiant Disorder |
| Other more distant relative(s) with ASD |
| Personality Disorder |
| Premature birth (delivery before 37 weeks) |
| Problems with eating foods - not diagnosed by a professional |
| Pyloric stenosis (blockage from stomach to small intestine) |
| RBS-r total score |
| Schizophrenia, Other Psychosis or Schizoaffective Disorder |
| SCQ total score |
| Seizure disorder or epilepsy |
| Separation Anxiety |
| Serious prenatal infection (for example, German measles) |
| Sex assigned at birth |
| Short stature |
| Sister with ASD |
| Sleep Disorder or sleep problem diagnosed by a professional |
| Sleep problems not diagnosed by a professional |
| Small head size (microcephaly) |
| Social (Pragmatic) Communication Disorder |
| Social Anxiety Disorder/Social Phobia |
| Speech and language, intellectual disability/cognitive Impairment, learning disability (LD) or other developmental delay or developmental disability |
| Speech articulation problems |
| Spina bifida/myelomeningocele (baby born with open spine, spinal cord outside of the body) |
| Spine deformity |
| Strabismus |
| Suspected cause of ASD - Birth or delivery complication |
| Suspected cause of ASD - Don't know |
| Suspected cause of ASD - Drug or alcohol exposure in pregnancy |
| Suspected cause of ASD - Environmental exposures |
| Suspected cause of ASD - Genetic causes |
| Suspected cause of ASD - Immunizations |
| Suspected cause of ASD - Infection in infancy or early childhood |
| Suspected cause of ASD - Other |
| Suspected cause of ASD - Other medical conditions |
| Suspected cause of ASD - Problems during pregnancy |
| Tourette Syndrome or Tic Disorder |
| Traumatic brain injury (hospitalized) |
| twin participating in SPARK |
| Urinary or genital birth defect |
| V. Sameness Behavior- Total Subscale Score (RBS-r) |
| VI. Restricted Behavior- Total Subscale Score (RBS-r) |
| Vision or hearing conditions |

| |
|---|
| Year at diagnosis |
| Year of birth |

**eTable 2**. Pairwise correlations between informative characteristics of the D1 group.
This large table could be accessed at https://github.com/danaiz/Data-driven-characterization-of-individuals-with-a-delayed-autism-diagnosis/blob/main/net_corr_p_1_fdr.xlsx

**eTable 3.** Pairwise correlations between informative characteristics of the D2 group.
This large table could be accessed at https://github.com/danaiz/Data-driven-characterization-of-individuals-with-a-delayed-autism-diagnosis/blob/main/net_corr_p_2_fdr.xlsx

**eTable 4.** Pairwise correlations between informative characteristics of the timely diagnosis group. This large table could be accessed at https://github.com/danaiz/Data-driven-characterization-of-individuals-with-a-delayed-autism-diagnosis/blob/main/net_corr_p_ed_fdr.xlsx

**eTable 5. Feature participation in pairwise correlation 4-cliques.** Pairwise correlations between any two features were calculated separately within the T, D1, and D2 subgroups, and modeled as a dense graph of feature nodes connected by correlation edges. Each graph was analyzed via 4-cliques. This table summarizes the percent of 4-cliques each feature belongs to, as a measure of its information flow.

| | T cliques | D1 cliques | D2 cliques |
|---|---|---|---|
| Motor delay | 6.349206 | 2.247191 | 4.895105 |
| Birth defects | 3.703704 | 0 | 4.195804 |
| Mood, depression, anxiety or OCD | 1.058201 | 3.370787 | 3.496503 |
| Oppositional defiant disorder | 0 | 0 | 3.030303 |
| Disruptive mood dysregulation disorder | 0 | 0 | 3.030303 |
| Depression or dysthymia | 0 | 1.123596 | 2.797203 |
| Cognitive impairment | 2.645503 | 1.123596 | 2.564103 |
| Gastrointestinal birth defects | 2.116402 | 0 | 2.564103 |
| Brain and spinal cord birth defects | 1.587302 | 0 | 2.564103 |
| DCDQ - fine motor handwriting | 0 | 2.247191 | 2.564103 |
| RBS-r - final score | 2.645503 | 3.370787 | 2.331002 |
| Social anxiety disorder or Social phobia | 0.529101 | 3.370787 | 2.331002 |
| speech | 0.529101 | 3.370787 | 2.331002 |
| Learning disability | 5.291005 | 3.370787 | 2.097902 |
| Birth defects of bones, hands or feet | 1.587302 | 0 | 2.097902 |
| Language delay or language disorder | 0.529101 | 3.370787 | 2.097902 |
| Spine deformity | 0.529101 | 0 | 2.097902 |
| RBS-r - compulsive behavior score | 0 | 1.123596 | 2.097902 |
| Growth conditions | 5.291005 | 0 | 1.864802 |
| DCDQ final score | 2.116402 | 4.494382 | 1.864802 |
| OCD | 1.058201 | 2.247191 | 1.864802 |
| Anxiety disorder | 0.529101 | 3.370787 | 1.864802 |
| Urinary or genital birth defect | 0.529101 | 0 | 1.864802 |
| RBS-r - ritualistic behavior | 0 | 3.370787 | 1.864802 |
| RBS-r - sameness | 0.529101 | 2.247191 | 1.631702 |
| Heart or lung birth defect | 0.529101 | 0 | 1.631702 |
| Separation anxiety | 0 | 1.123596 | 1.631702 |
| Attention or behavior disorders | 3.174603 | 0 | 1.398601 |
| Neurological conditions | 2.116402 | 0 | 1.398601 |
| Missing or malformed bones | 1.058201 | 0 | 1.398601 |
| Congenital heart disease or defect | 0.529101 | 0 | 1.398601 |
| RBS-r - restricted behavior | 0 | 5.617978 | 1.398601 |
| Intermittent explosive disorder | 0 | 0 | 1.398601 |
| Bipolar disorder | 0 | 0 | 1.398601 |
| Sleep disorder diagnosed by a professional | 5.820106 | 0 | 1.165501 |
| Premature birth (delivery before 37 weeks) | 2.645503 | 3.370787 | 1.165501 |

| | | | |
|---|---|---|---|
| **Speech articulation problems** | 1.587302 | 2.247191 | 1.165501 |
| **Brain malformation or abnormality (shown on MRI)** | 0.529101 | 0 | 1.165501 |
| **Lung malformation** | 0.529101 | 0 | 1.165501 |
| **Seizure disorder or epilepsy** | 0 | 0 | 1.165501 |
| **Intellectual , cognitive , global** | 3.703704 | 1.123596 | 0.932401 |
| **Vision or hearing conditions** | 3.174603 | 0 | 0.932401 |
| **Difficulty gaining weight** | 2.116402 | 0 | 0.932401 |
| **RBS-r stereotyped behavior score** | 0.529101 | 5.617978 | 0.932401 |
| **Conduct disorder** | 0 | 0 | 0.932401 |
| **SCQ score** | 3.174603 | 5.617978 | 0.699301 |
| **Year of birth** | 2.645503 | 2.247191 | 0.699301 |
| **Birth or pregnancy complications** | 2.116402 | 2.247191 | 0.699301 |
| **Cognitive delays or impairment due to a medical condition or exposure** | 1.058201 | 0 | 0.699301 |
| **Facial birth defect** | 1.058201 | 0 | 0.699301 |
| **RBS-R - self injurious behavior** | 0.529101 | 1.123596 | 0.699301 |
| **Short stature** | 0.529101 | 0 | 0.699301 |
| **Hoarding** | 0 | 0 | 0.699301 |
| **Twin participating in SPARK** | 2.645503 | 3.370787 | 0.4662 |
| **Twin** | 2.645503 | 3.370787 | 0.4662 |
| **No twin** | 2.645503 | 3.370787 | 0.4662 |
| **DCDQ - control during movement** | 2.116402 | 2.247191 | 0.4662 |
| **Insufficient oxygen at birth with NICU stay** | 1.587302 | 1.123596 | 0.4662 |
| **ADHD** | 0.529101 | 0 | 0.4662 |
| **Intraventricular hemorrhage at birth** | 0.529101 | 0 | 0.4662 |
| **DCDQ - general coordination** | 0 | 4.494382 | 0.4662 |
| **Strabismus** | 0 | 0 | 0.4662 |
| **Pyloric stenosis** | 0 | 0 | 0.4662 |
| **Extra fingers and or or extra toes** | 0 | 0 | 0.4662 |
| **Missing kidney** | 0 | 0 | 0.4662 |
| **Monozygotic twin** | 1.058201 | 2.247191 | 0.2331 |
| **Social communication disorder** | 1.058201 | 1.123596 | 0.2331 |
| **Suspected cause of ASD - Birth or delivery complication** | 1.058201 | 1.123596 | 0.2331 |
| **Dizygotic twin** | 1.058201 | 1.123596 | 0.2331 |
| **Suspected cause of ASD - Problems during pregnancy** | 0.529101 | 1.123596 | 0.2331 |
| **Macrocephaly** | 0 | 0 | 0.2331 |
| **Intestinal malrotation** | 0 | 0 | 0.2331 |
| **Cleft palate** | 0 | 0 | 0.2331 |
| **Microcephaly** | 0 | 0 | 0.2331 |
| **Feeding or eating problems** | 3.174603 | 0 | 0 |
| **Nuclear family ASD** | 1.058201 | 1.123596 | 0 |

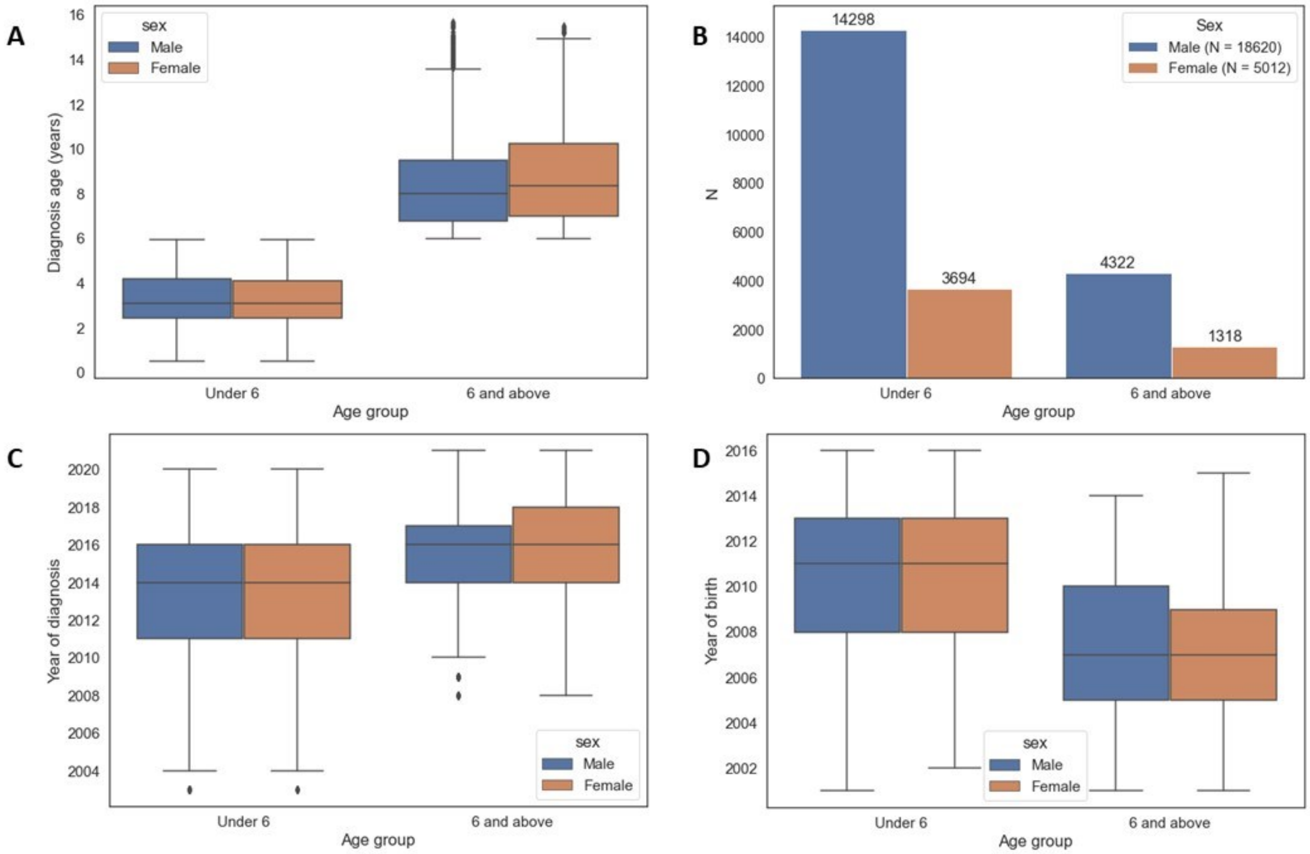


**eFigure 1**. Sex and age of ASD diagnosis of SPARK individuals included in this study. (**A**) The distribution of age at diagnosis by sex; (**B**) The number of individuals in each group; (**C**) The distribution of years in which the diagnosis was received; (**D**) Year of birth distribution by age group and sex.

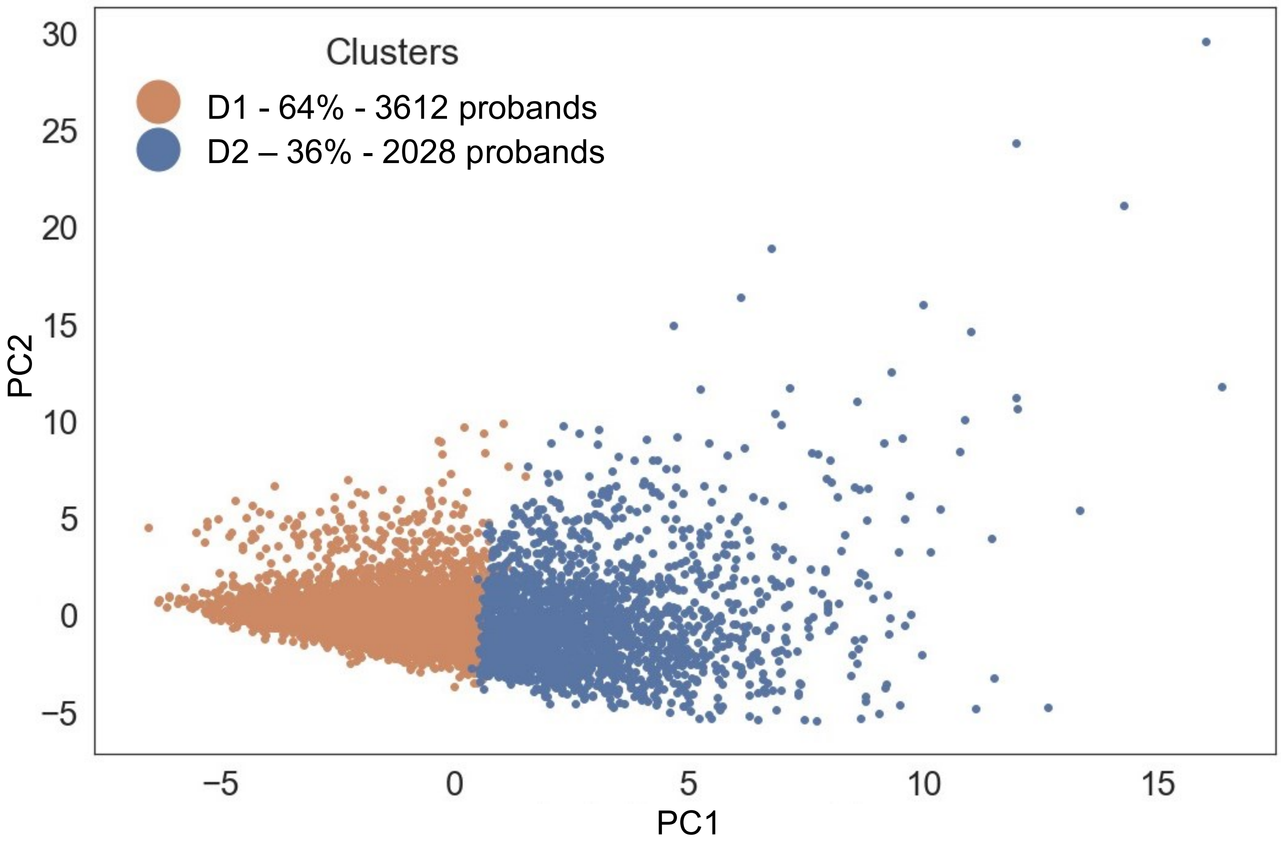


**eFigure 2**. Two subtypes of individuals with delayed autism diagnosis. Shown are the first two principal components of all variables listed in eTable 1.

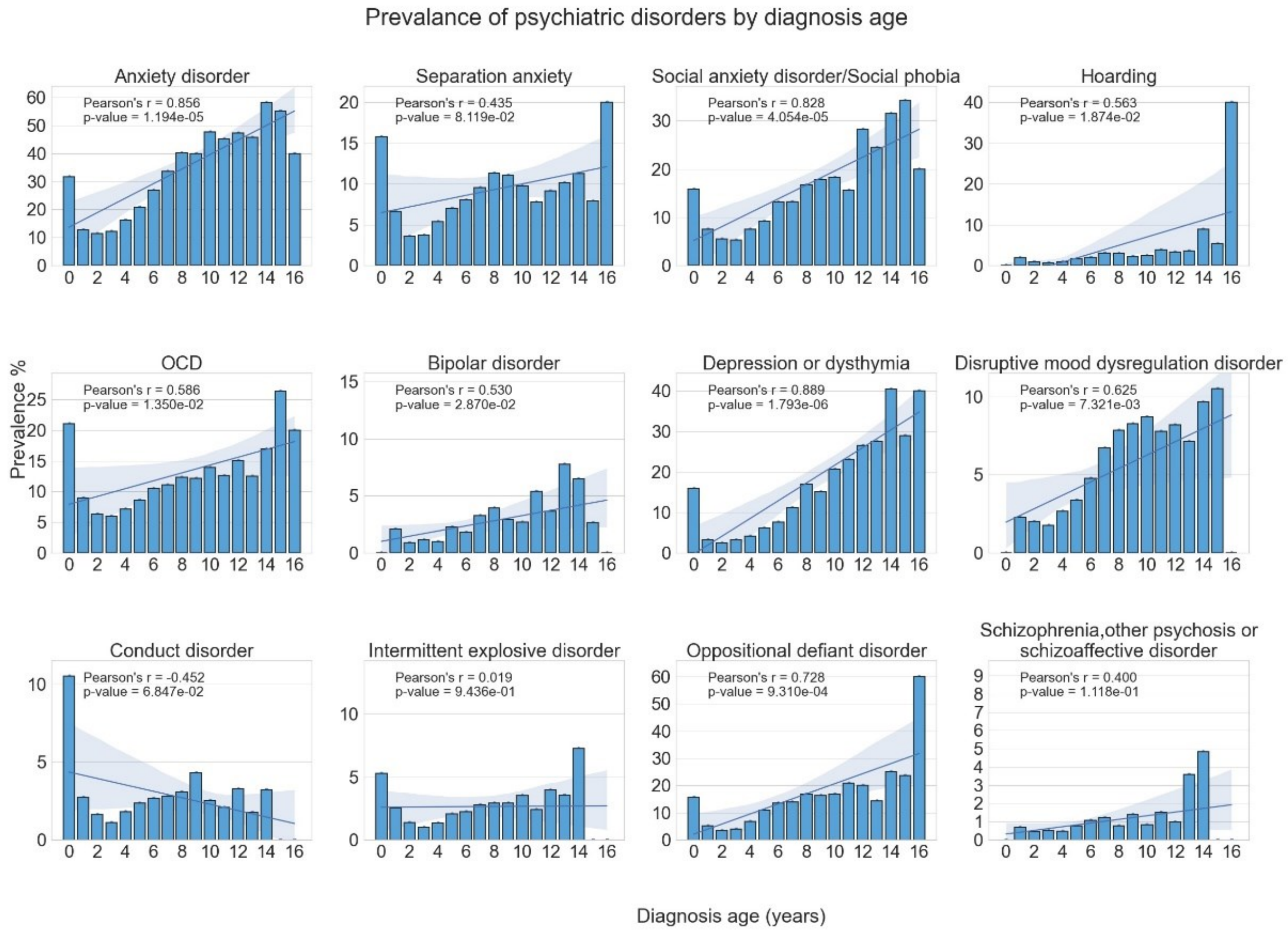


**eFigure 3.** Prevalence of psychiatric disorders according to the age of autism diagnosis. For each disorder, Pearson's correlation was calculated between the age of autism diagnosis and the prevalence of the disorder in individuals diagnosed at that age. The least squares regression line is shown along its 95% confidence interval.

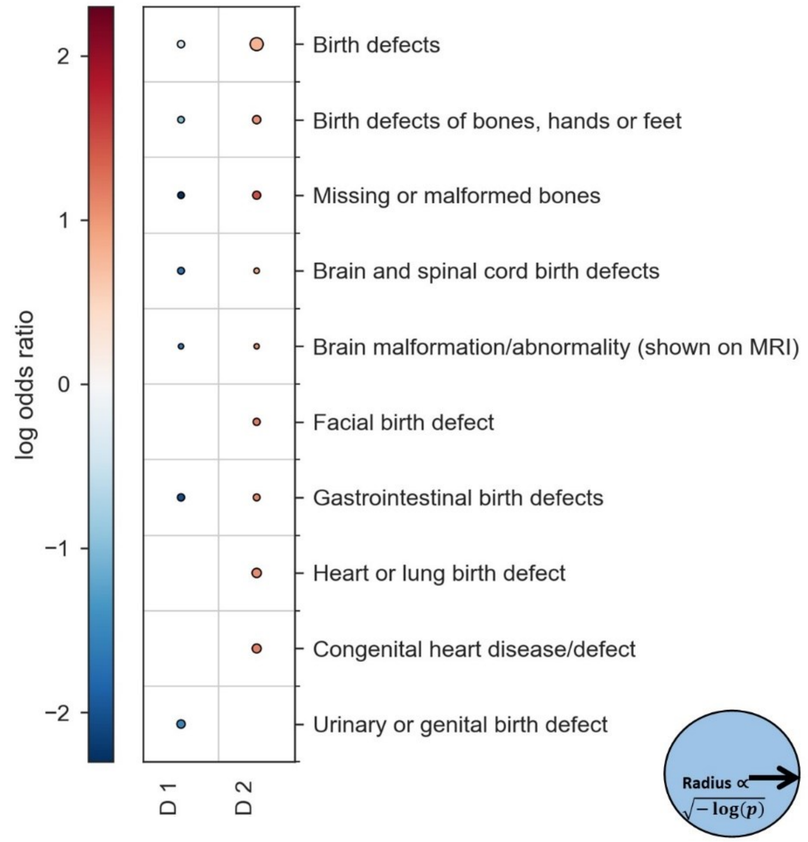


**eFigure 4**. Differences in congenital disorders between individuals in each delayed diagnosis group and those that received a timely diagnosis. The color of each circle represents the effect size measured by the log ratio of a feature's mean in each group as compared to the feature's mean among timely diagnosed individuals. The size of the circles is proportional to $\sqrt{-log(P\ value)}$. Thus, the larger the circle, the smaller the P value. The lowest possible P value in this analysis is $6.87 \times 10^{-278}$. Circles are shown only for significant results.

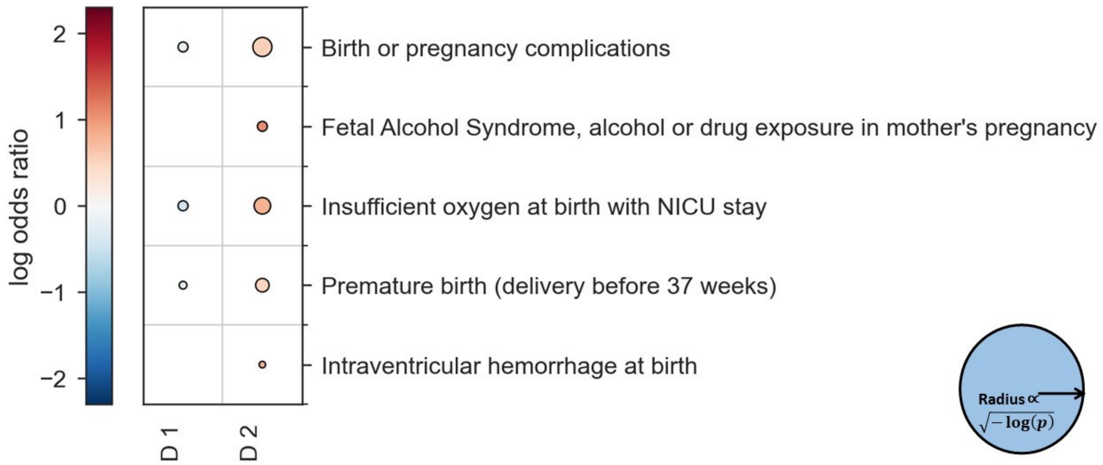


**eFigure 5**. Differences in prenatal and perinatal complications between individuals in each delayed diagnosis group and those with a timely diagnosis. The color of each circle represents the effect size measured by the log ratio of a feature's mean in each group as compared to the feature's mean among timely diagnosed individuals. The size of the circles is proportional to $\sqrt{-log(P\ value)}$. Thus, the larger the circle, the smaller the P value. The lowest possible P value in this analysis is $6.87 \times 10^{-278}$. Circles are shown only for significant results.

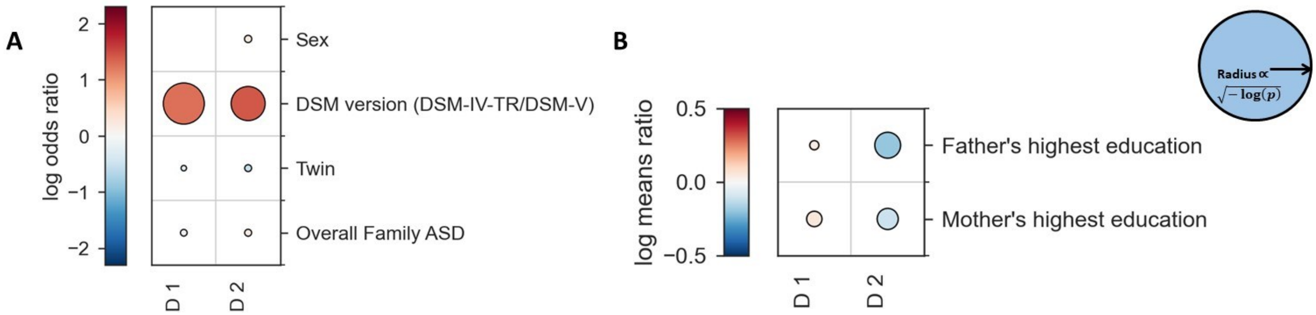


**eFigure 6**. Differences in family and demographics between individuals in each delayed diagnosis group and those with a timely autism diagnosis. The color of each circle represents the effect size measured by the log ratio of a feature's mean in each group as compared to the feature's mean among timely diagnosed individuals. The size of the circles is proportional to $\sqrt{-log(P\ value)}$. Thus, the larger the circle, the smaller the P value. The lowest possible P value in this analysis is $6.87 \times 10^{-278}$. Circles are shown only for significant results.

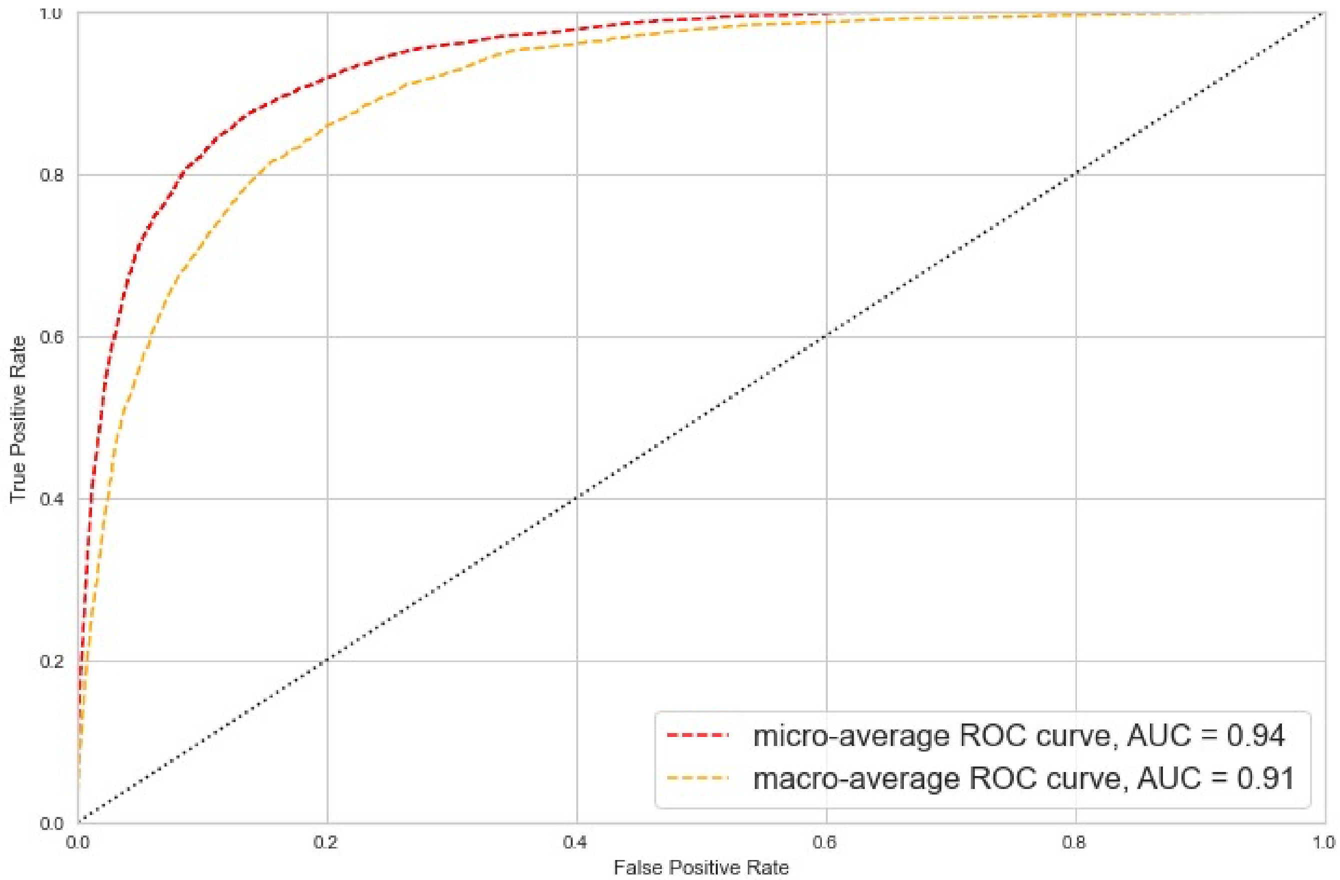


**eFigure 7.** Receiver operating characteristics (ROC) curve for distinguishing the two delayed diagnosis groups using a random forest classifier.

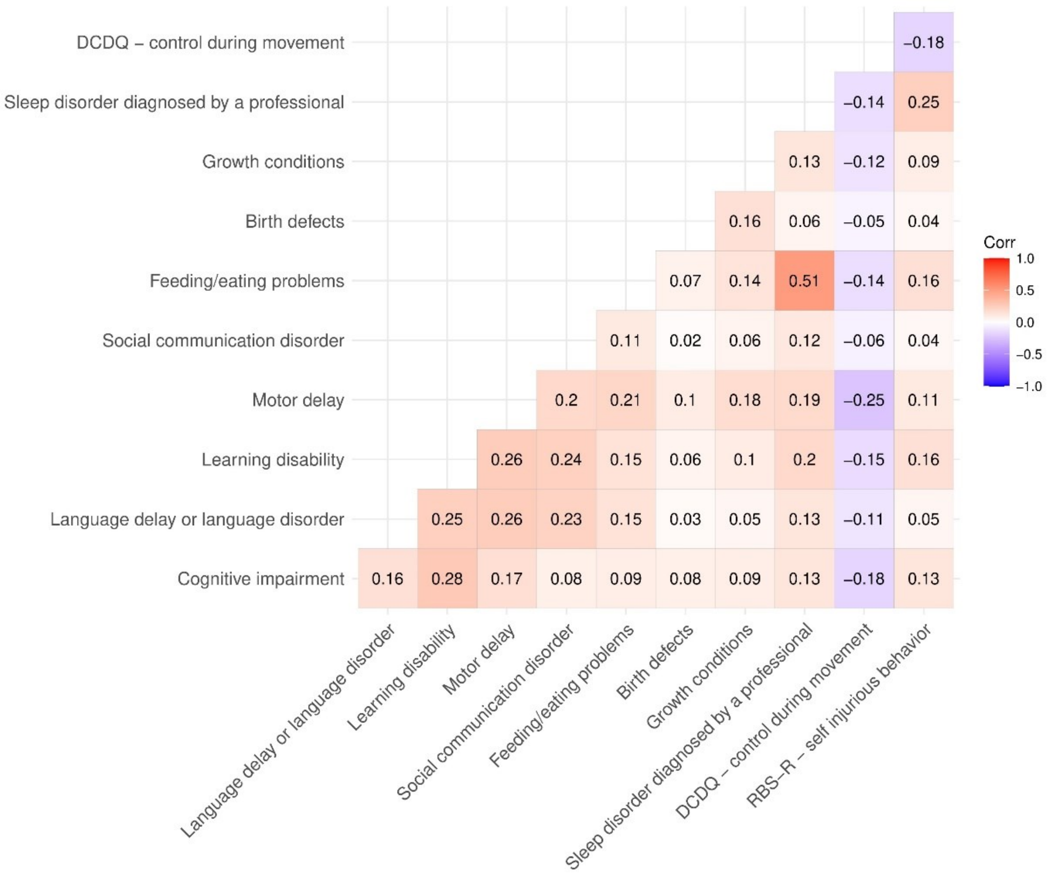


**eFigure 8.** Heatmap of variables most strongly correlated with the top connected features of the timely diagnosis group.
